# Waning immunity of the BNT162b2 vaccine: A nationwide study from Israel

**DOI:** 10.1101/2021.08.24.21262423

**Authors:** Yair Goldberg, Micha Mandel, Yinon M. Bar-On, Omri Bodenheimer, Laurence Freedman, Eric J. Haas, Ron Milo, Sharon Alroy-Preis, Nachman Ash, Amit Huppert

## Abstract

**Background:** Starting December 2020, Israel began a mass vaccination campaign against coronavirus administering the Pfizer BNT162b2 vaccine, which led to a sharp curtailing of the outbreak. After a period with almost no SARS-CoV-2 infections, a resurgent COVID-19 outbreak initiated mid June 2021. Possible reasons for the breakthrough were reduced vaccine effectiveness against the Delta variant, and waning immunity. The aim of this study was to quantify the extent of waning immunity using Israel’s national-database.

**Methods:** Data on all PCR positive test results between July 11-31, 2021 of Israeli residents who became fully vaccinated before June 2021 were used in this analysis. Infection rates and severe COVID-19 outcomes were compared between individuals who were vaccinated in different time periods using a Poisson regression, stratifying by age group and adjusting for possible confounding factors.

**Results:** The rates of both documented SARS-CoV-2 infections and severe COVID-19 exhibit a statistically significant increase as time from second vaccine dose elapsed. Elderly individuals (60+) who received their second dose in March 2021 were 1.6 (CI: [1.3, 2]) times more protected against infection and 1.7 (CI: [1.0, 2.7]) times more protected against severe COVID-19 compared to those who received their second dose in January 2021. Similar results were found for different age groups.

**Conclusions:** These results indicate a strong effect of waning immunity in all age groups after six months. Quantifying the effect of waning immunity on vaccine effectiveness is critical for policy makers worldwide facing the dilemma of administering booster vaccinations.

## INTRODUCTION

A key to the management of the COVID-19 pandemic is mass vaccination of the population. Yet, the success of this policy is challenged by the possibility of breakthrough infection and disease in fully vaccinated persons. One potential cause of breakthrough is the emergence of new variants of concern (VOC)^1^ that escape immunity, reducing the effectiveness of the vaccine in preventing disease and viral transmission. Some studies published on Pfizer BNT162b2 vaccine effectiveness against the Beta^2,3^ and Delta^2,4,5,6^ variants showed only modest levels of breakthrough infection and disease while others showed higher rates ^7,8^. A second potential cause of breakthrough is waning of the immunity conferred by the vaccine. Mass vaccination with the BNT162b2 vaccine began in December 2020, so little is currently known about waning immunity in the medium and long term. However, a recent paper on longer-term follow-up of participants in the Phase 2/3 randomized trial of the BNT162b2 vaccine^9^ reported a reduction in vaccine efficacy from 96% (7d to <2m) to 90% (2m to <4m) to 84% (4m to ∼7m). There is also a preliminary report of waning effectiveness of the same vaccine from a Health Maintenance Organization in Israel^10^, and evidence of a decay in vaccine-induced neutralization titers during the first six months following the second dose^11^.

Israel conducted a very successful vaccination campaign using the BNT162b2 vaccine ^12,13,14^. Starting in December 2020, ∼5,860,000 individuals were vaccinated and ∼12,400,000 doses administered, leading to a sharp curtailing of the outbreak. By May 2021, infection rates dropped more than a hundred-fold from the peak values to a few dozen weekly cases, most of which were unvaccinated individuals or people returning from abroad. However, the number of positive polymerase chain reaction (PCR) tests started to rise exponentially (doubling roughly every 10 days) during June 2021, with a significant number of infections reported in vaccinated individuals (Figure 1). This rise in community transmission was followed by a concomitant rise in the number of severe cases and deaths, in both the vaccinated and unvaccinated population. Genetic analysis based on whole genome sequencing revealed that as of June 2021, more than 98% of positive cases in Israel are attributed to the Delta variant^15^.

**Figure 1:**
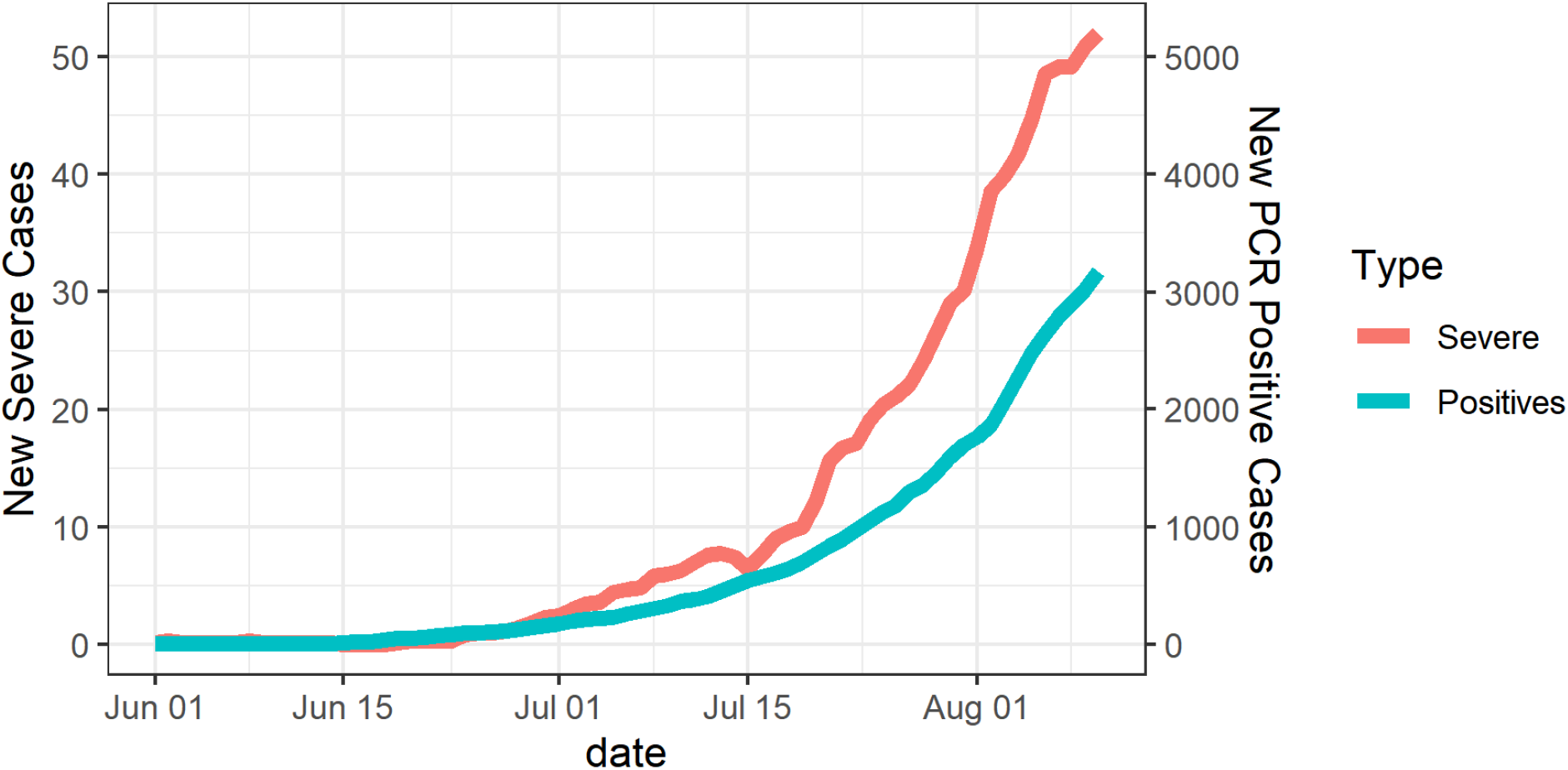
Number of new documented SARS-CoV-2 infections and new severe COVID-19 in the studied population during the Delta wave in Israel.

Here, we estimate the extent of immune evasion by the Delta variant and the role of waning immunity in the observed breakthrough.

## METHODS

### Data

Data on all residents of Israel who became fully vaccinated before June 1st, 2021 and who were not infected before the study period were extracted from the Ministry of Health database on August 10, 2021. Data include vaccination dates (first and second doses), PCR tests (all dates and results), hospitalization admission date (if relevant), clinical severity status (hospitalization, severe, death) and demographic variables such as age, gender, and demographic group (General Jewish, Arab, ultra-Orthodox Jews).

We extracted from the database all documented SARS-CoV-2 infections diagnosed in the period in which the Delta variant was dominant, and the severity of the disease following infection. The Delta variant became dominant in Israel during June 2021, after several weeks of almost no new cases in the country. We focus on infections documented between July 11-31, 2021 (inclusive; study period), removing from the data all confirmed cases documented before that period. The start date was chosen as a time when the virus had already spread throughout the entire country and across demographic groups. As the disease is mild and often asymptomatic in young people, we omit from all analyses children 16 years or younger (most of them were unvaccinated or were vaccinated for only a short time), and for several analyses we consider only persons aged 40 years or older as severe disease was rare in the younger population.

During the study period, about 10% of infections were detected in residents of Israel returning from abroad. These infections were acquired outside Israel and were less likely to be of the Delta variant, although not all specimens underwent whole genome sequencing (WGS). Moreover, most residents who traveled abroad were vaccinated and were exposed to different populations, hence their risk of infection differs from that of people in Israel. We therefore removed from the main analysis all residents returning from abroad during July. Figure 2 reports the pre-processing of the data. Out of 5,223,680 fully vaccinated adults, we retained 4,785,245 individuals for the main analysis.

**Figure 2:**
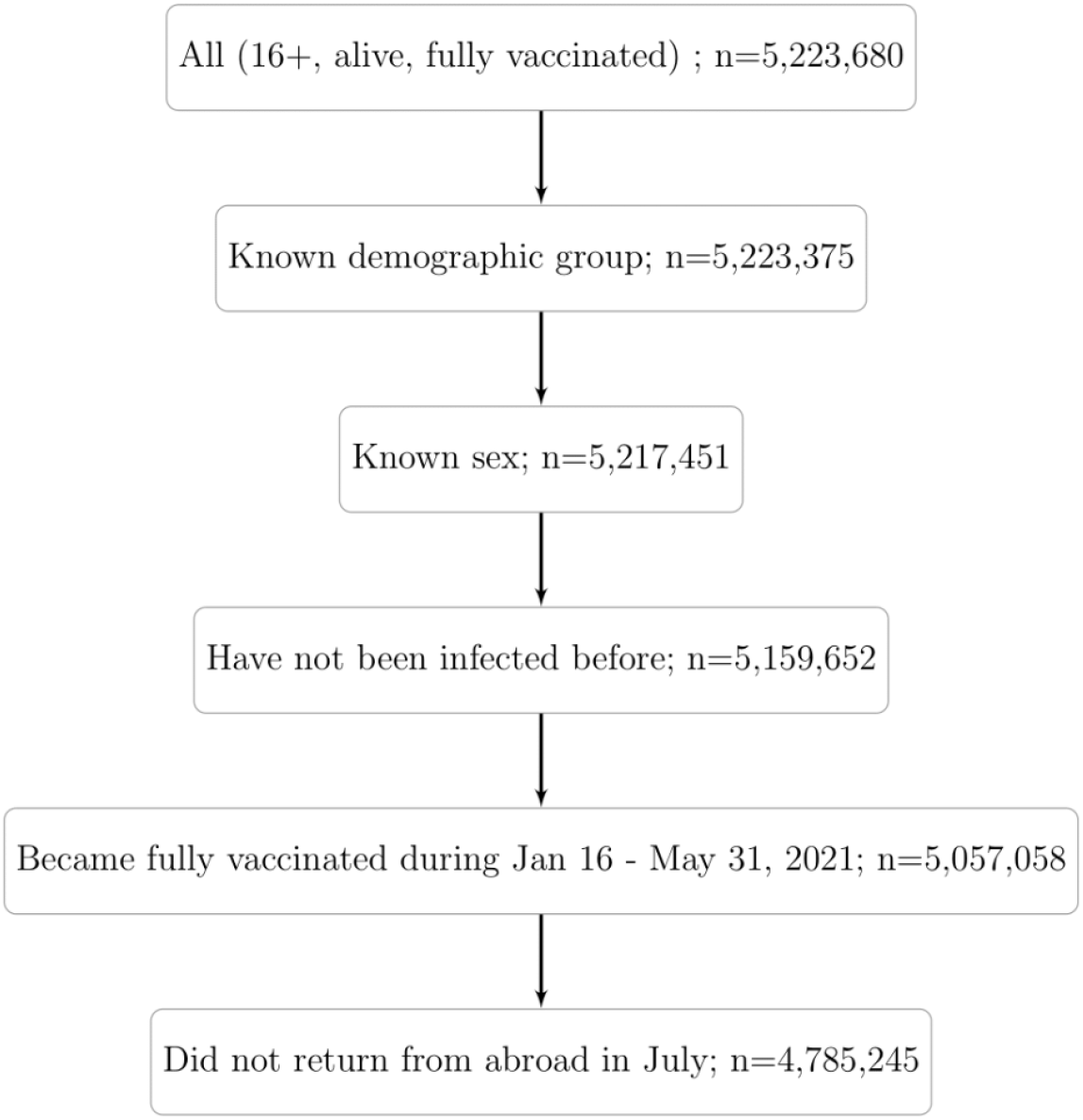
Study population. The population includes people who were fully vaccinated prior to June 1, 2021, were not abroad during July 2021, and had no documented SARS-CoV-2 PCR-positive result before July 11, 2021.

### Statistical Analysis

The vaccination regimen in Israel was such that the second dose was administered three weeks after the first dose in the vast majority of recipients. We defined fully vaccinated individuals as those for which seven days or more have passed since receiving the second BNT162b2 dose. All residents aged 60 years or older were eligible for vaccination at no charge starting December 20, 2020, thus becoming fully vaccinated starting mid-January 2021. At that time, younger people were eligible for vaccination only if they belonged to a risk group (e.g., healthcare workers or people having one of several chronic diseases). On January 12, 2021, the eligibility age was reduced to 55 years old, and then on January 19, 2021, to 40 years old. On February 4th, all individuals 16 and older were eligible for vaccination. Thus, if not belonging to a risk group, people aged 40-59 and 16-39 received the second dose starting mid-February and beginning of March, respectively. Based on these dates, we defined our periods of interest in half months starting from January 16; the vaccination period of an individual is determined by the time he/she became fully vaccinated. All analyses were stratified by vaccination period and age group (16-39, 40-59, and 60+).

The association between infection rate and period of vaccination is a measure of waning immunity. Indeed, provided that proper controlling for confounding variables is done, without immunity waning, we would expect to see no differences in infection rates between persons vaccinated at different times. In order to examine the effect of waning immunity during a period when the Delta variant was predominant, we compared the infection rate (per 1,000 people) during the study period (July 11-31) among individuals who were fully vaccinated in different periods. Confidence intervals for the rates (95%), based on standard confidence intervals for proportions, were calculated. A similar analysis was performed to compare the association between vaccination period and severe COVID-19, but for this outcome we used a period of a whole month, as there were few cases of severe disease.

In order to account for possible confounders, we fitted Poisson regressions. The outcome variable is the number of documented SARS-CoV-2 infections (or severe COVID-19) during the study. The period of vaccination, seven days after the second dose of COVID-19 vaccine, is the primary exposure of interest. The models compare the protection between different vaccination periods, where the earliest period is set as the reference group to which all other periods are compared. A different effect of the vaccination period for each age group was allowed by including an interaction term between age and vaccination period. Additional potential confounders were added as covariates, as described below. All analyses were performed using the glm function in the R Statistical Software^10^ with the natural logarithm of the number of individuals in each group added as an offset.

In addition to age and gender, the regression analysis was adjusted for the following confounders. First, as the number of new events rose rapidly during the study (Figure 1), we included the week in which the infection was first documented in the analysis. Second, although PCR testing is free in Israel for all residents, some people tend to comply more with medical recommendations including likelihood to be tested than others. This can affect the probability of performing PCR tests, hence it is a possible source of detection bias. To partially account for this, we stratified individuals according to the number of PCR tests they underwent before the vaccination campaign started, during the period March 1, 2020 - November 31, 2020. We define three risk levels: no past PCR tests, one test, and two or more tests. Finally, there are three major demographic groups in Israel with very different cultures and varying compliance with medical recommendations. The proportion of vaccinated individuals, as well as exposure to the virus, differ among these groups^16^. Although we restricted the study to dates when the virus was widespread throughout the entire country, we added as a covariate an indicator for demographic group (General Jewish, Arab, ultra-Orthodox) in order to eliminate the residual effect of this variable on the results.

## RESULTS

The study data describes 4,785,245 individuals. Of these individuals, 12,927 had a positive PCR test and 348 deteriorated to a severe condition. Table 1 gives the number of events by vaccination period, and Table S1 in the Supplementary Appendix provides a more detailed summary by vaccination period and age group. Table 1 also compares the characteristics of individuals by vaccination period. Due to the risk-based vaccination policy, people who were vaccinated in January were older than those who were vaccinated later on. In addition, the lower risk of COVID-19 related complications in younger people may have caused people to believe that vaccination was not urgent or even necessary, which also affected the age distribution over the months. The distribution of the number of past PCR tests (tests taken before vaccination started) changed slightly between the periods, with 65% having no previous tests in the 2^nd^ half of January compared to 75% in May. There is a significant difference in time of vaccination between the main demographic groups, where Arabs and ultra-Orthodox Jews received vaccines later than the general Jewish population. This is partially because these groups are younger than the general population, but mainly because of cultural differences^16^.

**Table 1:** Demographic and clinical characteristics of the study population for the different vaccination periods. A and B indicate the first and second halves of the corresponding months.

| Variable (%) | Jan 16-31 | Feb 1-15 | Feb 16-28 | Mar 1-15 | Mar 16-31 | Apr | May |
| --- | --- | --- | --- | --- | --- | --- | --- |
| Total vaccinated (n) | 1,073,766 | 971,218 | 746,944 | 818,975 | 748,932 | 324,996 | 100,414 |
| Positive SARS-CoV-2 PCR tests | 3,640 | 3,073 | 2,182 | 2,065 | 1,383 | 443 | 141 |
| Severe COVID-19 | 215 | 95 | 12 | 16 | 5 | 5 | 0 |
| gender = male | 516,841 (48%) | 458,529 (47%) | 379,708 (51%) | 410,413 (50%) | 358,155 (48%) | 153,600 (47%) | 46,426 (46%) |
| Age: 16-39 | 125,587 (12%) | 195,580 (20%) | 352,247 (47%) | 549,089 (67%) | 496,455 (66%) | 217,622 (67%) | 67,281 (67%) |
| Age: 40-59 | 242,866 (23%) | 417,618 (43%) | 327,766 (44%) | 208,072 (25%) | 190,200 (25%) | 78,249 (24%) | 22,240 (22%) |
| Age: 60+ | 705,313 (66%) | 358,020 (37%) | 66,931 (9%) | 61,814 (8%) | 62,277 (8%) | 29,125 (9%) | 10,893 (11%) |
| Previous SARS-CoV-2 PCR tests = 0 | 698,648 (65%) | 653,978 (67%) | 500,610 (67%) | 564,927 (69%) | 536,538 (72%) | 240,330 (74%) | 75,668 (75%) |
| Previous SARS-CoV-2 PCR tests = 1 | 203,763 (19%) | 196,859 (20%) | 163,873 (22%) | 172,492 (21%) | 144,029 (19%) | 56,872 (18%) | 16,345 (16%) |
| Previous SARS-CoV-2 PCR tests = 2+ | 171,355 (16%) | 120,381 (12%) | 82,461 (11%) | 81,556 (10%) | 68,365 (9%) | 27,794 (9%) | 8,401 (8%) |
| Demographic: General Jewish Population | 968,286 (90%) | 825,615 (85%) | 616,653 (83%) | 657,238 (80%) | 50,6777 (68%) | 20,1970 (62%) | 72,400 (72%) |
| Demographic: ultra-Orthodox Jews | 43,841 (4%) | 38,309 (4%) | 40,371 (5%) | 46,801 (6%) | 44,427 (6%) | 20,546 (6%) | 7,363 (7%) |
| Demographic: Arabs | 61,639 (5%) | 107,294 (11%) | 89,920 (12%) | 114,936 (14%) | 197,728 (26%) | 102,480 (32%) | 20,651 (21%) |

The rate of documented SARS-CoV-2 infections exhibits a clear increase as a function of time from vaccination (Figure 3). For example, among people 60 years or older who were fully vaccinated in the 2nd half of January, the rate is 3.2 cases for 1,000 persons, compared to 2.1 and 1.6 for those who were vaccinated in the 2nd halves of February and March, respectively (Figure 3). Similar results are shown for people in the other age groups (Figure 3) and when categorizing age into decades (Figure S1 in the Supplementary Appendix). However, for age groups 16-39 and 40-59, mainly healthcare workers and people at higher risk of infection were vaccinated during the first two and three periods, respectively, thus, the first bars of these groups may be biased due to selective samples and should be interpreted with care.

**Figure 3:**
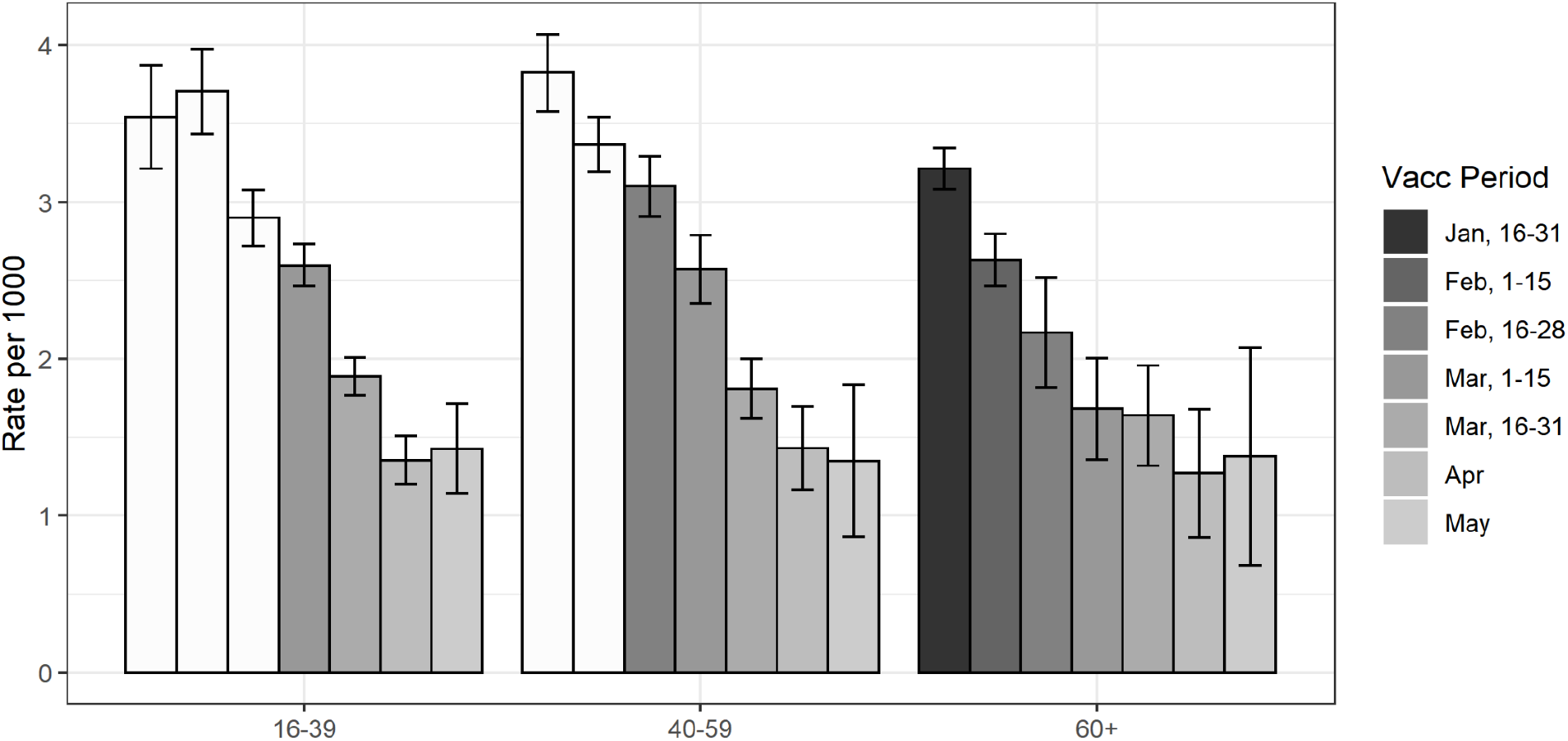
Rate of documented SARS-CoV-2 infection (per 1,000 persons) from July 11, 2021 to July 31, 2021, stratified by period of second dose of COVID-19 vaccine and age group. White bars represent periods at which only persons at higher risk were allowed to receive vaccination.

A similar pattern is obtained when analyzing severe disease for the older (60+) group (Figure 4), in which vaccination periods are defined as full months due to small group size. The rate of severe cases for people aged 60 or older who were fully vaccinated in January is 0.29 for 1000 persons, and is reduced to 0.23, 0.15, and 0.10 for those who were fully vaccinated in February, March, and April-May, respectively. The numbers of severe cases in the younger age groups are too small to draw conclusions.

**Figure 4:**
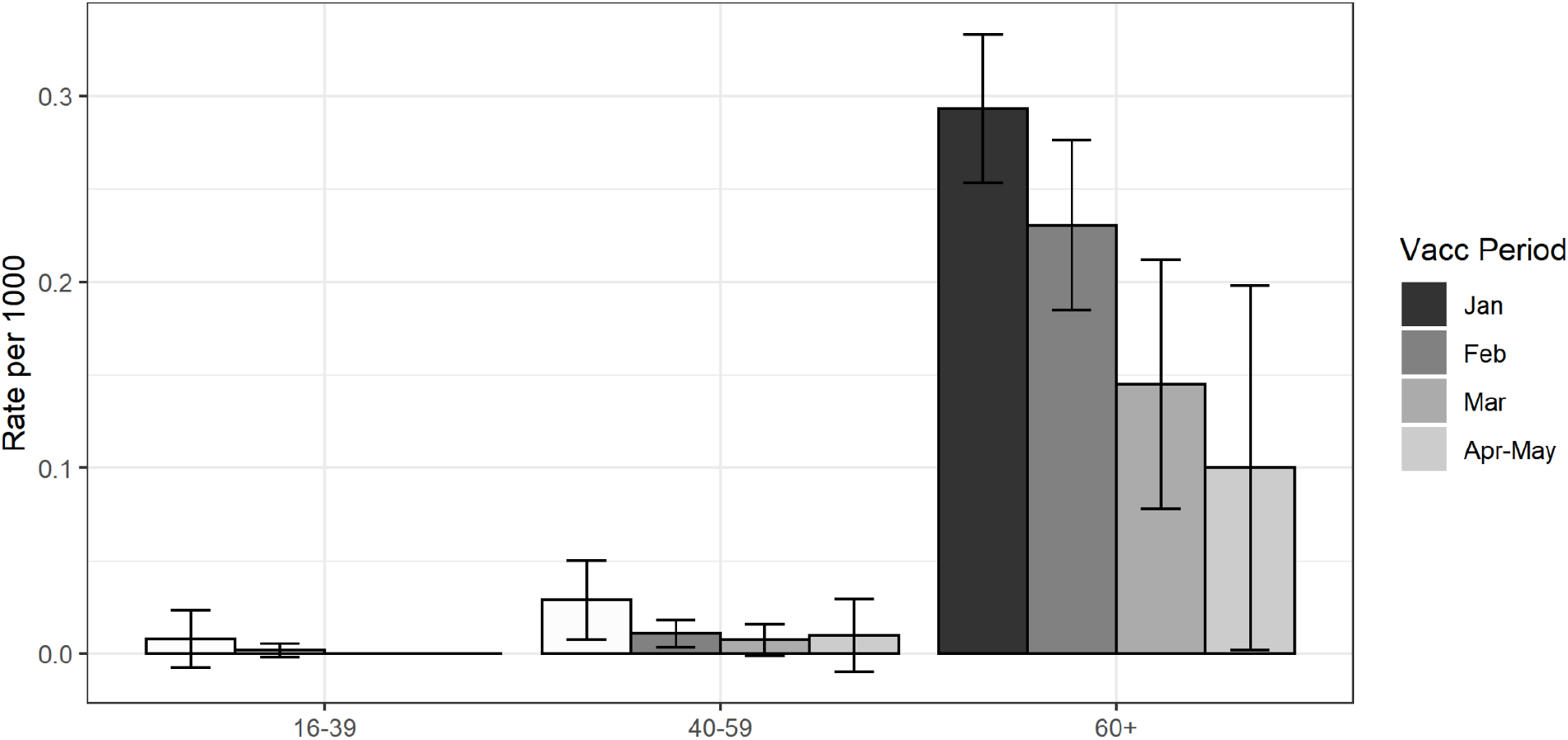
Rate of severe COVID-19 (per 1,000 persons) from July 11, 2021 to July 31, 2021, stratified by period of second dose of COVID-19 vaccine and age group. White bars represent periods at which only persons at higher risk were allowed to receive vaccination.

Table 2 presents the regression results for the documented SARS-CoV-2 infection and severe COVID-19 outcomes; the complete set of parameters is provided in Tables S2 and S3 of the Supplementary Appendix. The numbers in the tables are measures of protection given by the ratio between the estimated rate in the first period (second half of January) and the other periods. For the age group 60 and over, the protection against documented infection for those vaccinated in the first half of February is 1.1 compared to those vaccinated in the second half of January. The protection further increases to 1.6 in March and to 2.1 in April-May. The same phenomenon, of increased protection with decreased time from vaccination, is observed in the other two age groups. There are fewer severe cases in people younger than 60, especially in the 16-39 group (Table S1, Supplementary Appendix), so the model could be fitted only to the 40-59 and 60+ groups and only for the months January-March. As can be observed, the confidence intervals are wide, but the results suggest a monotonic decrease in protection against severe disease over time.

**Table 2:** Protection against documented SARS-CoV-2 infection and severe COVID-19 [95% CI] compared to the first period (January 16-31) for the different age groups, adjusted for week of infection, past PCR testing (none, one or 2+), demographic groups, and gender. For severe COVID-19, estimates are not provided for the lowest age group and for the last vaccination periods due to very low case numbers. A and B indicate the first and second halves of the corresponding months.

| OUTCOME = Positive SARS-CoV-2 PCR test |  |  |  |  |  |  |
| --- | --- | --- | --- | --- | --- | --- |
| Age | FebA | FebB | MarA | MarB | Apr | May |
| 16-39 | 0.9 [0.8, 1] | 1.2 [1, 1.3] | 1.3 [1.1, 1.4] | 1.5 [1.4, 1.7] | 2 [1.7, 2.3] | 2 [1.6, 2.5] |
| 40-59 | 1.1 [1, 1.1] | 1.1 [1, 1.2] | 1.2 [1.1, 1.4] | 1.6 [1.4, 1.8] | 1.9 [1.6, 2.4] | 2.3 [1.6, 3.3] |
| 60+ | 1.1 [1.1, 1.2] | 1.3 [1.1, 1.5] | 1.6 [1.3, 2] | 1.6 [1.3, 2] | 2.1 [1.5, 2.9] | 2.1 [1.2, 3.4] |
| OUTCOME = Severe COVID-19 |  |  |  |  |  |  |
| Age | Feb | Mar |  |  |  |  |
| 40-59 | 2.2 [0.8, 6.1] | 2.8 [0.7, 10.9] |  |  |  |  |
| 60+ | 1.2 [0.9, 1.5] | 1.7 [1.0, 2.7] |  |  |  |  |

## DISCUSSION

Israel has a centralized healthcare system and succeeded in vaccinating most of its population in a very short period of time^10,11,12^. It is therefore a unique population for studying the effects of the BNT162b2 vaccine on the spread and severity of COVID-19, as well as waning of vaccine protection with time. The appearance and rapid predominance of the Delta variant during June 2021 resulted in a dramatic increase in the number of new SARS-CoV-2 infections in people who were fully vaccinated, which raised the question of decreased efficacy of the vaccine over time (Figure 1). A comparison of the rate of infection (per 1000 persons) among people who were vaccinated at different times reveals a clear reduction in protection in all age groups, with and without correction for measured confounding factors (Figure 3 and Table 2). For PCR confirmed infections, the protection for people aged 60 and over who were vaccinated in March is 1.5 higher than the protection for individuals in the same age group who were vaccinated in the second half of January. The data shows similar reduction in protection for other age groups. The data also exhibits reduction in protection against severe COVID-19 as a function of time from vaccination. Serological studies in Israel show time-dependent reduction in neutralization titers^11,17^, which further supports the finding in this population-based research.

In contrast to early findings from the United Kingdom (UK)^6^, in Israel about two thirds of severe COVID-19 during the study period were among individuals who received two doses of the BNT162b2 vaccine. Two major differences exist between the vaccination policies of Israel and the UK. First, the current analysis used data from July 2021, a time when for the majority of the Israeli population at least five months passed from the second dose to the outbreak of the Delta variant. The UK data were collected during April-June 2021 with a much shorter time from vaccination to the outbreak. Second, Israel has followed the original Pfizer protocol of administering the second dose three weeks (21 days) after the initial vaccination in the vast majority of recipients, while in the UK the time between doses has been typically longer^5^.

While a study of the vaccinated population is of most interest in terms of waning immunity, a comparison to unvaccinated individuals is of interest in order to predict future burden on the health system. The unvaccinated group above age 16 in Israel comprises people who could not get the vaccine due to contraindications or precautions, or people who hesitated to take the vaccine for various reasons. Data on unvaccinated individuals were not directly available, as some residents had not used the medical system during the pandemic and were not reflected in PCR testing or hospital databases. Moreover, people who were not vaccinated might differ from the vaccinated population in important characteristics that could result in biased estimates. Nevertheless, we estimated that the efficacy of the vaccine against documented infection for people aged 60 or older decreases from 73% for those who became fully vaccinated in the second half of March to 57% for those who became fully vaccinated during the second half of January; see Supplementary Appendix (Supplementary Analysis 1). A similar decrease in vaccine protection is observed for the other age groups. The efficacy of the vaccine against severe disease for the 60+ age group also decreases; from 91% to 86% between those vaccinated four months to those vaccinated six months before the study. The corresponding efficacies for the 40-59 age group are 98% and 94%. Thus, the vaccine seems to be highly effective even after six months compared to the unvaccinated population, but its effectiveness is significantly lower than it was closer to the vaccination date. This finding is in line with the findings from the randomized trial of Pfizer that reported a reduction in vaccine efficacy against symptomatic infection from 96% in the first two months after vaccination to 84% four to seven months after vaccination when averaged over all age groups combined^9^.

Observational studies are often subject to confounding bias and detection bias. The main risk factors for SARS-CoV-2 infection are patient characteristics and exposure risk. We adjusted for the former by including covariates that are known to be associated with infection, enabling the effect of age to vary among vaccination periods. Adjusting for exposure is more complicated. We therefore started our study period on July 11, 2021, a time when the disease was widespread throughout the country. Risk of exposure differs in Israel between demographic groups, which were controlled in the model. In addition, we fitted the models to the general Jewish population (which had the highest rates of breakthrough infection and disease), excluding ultra-Orthodox Jews and Arabs, and obtained similar results (data not shown). Finally, we added the week of infection as an additional covariate to the model in order to correct for the exponential growth of infection rate and exposure over time.

Eliminating detection bias is more difficult. As not all SARS-CoV-2 infections are detected and a PCR test is required to confirm infection, fewer infections may be confirmed in sub-groups that perform fewer tests. On the other hand, vaccinated individuals tend to test less, unless exposed to an infected person or when symptoms appear. Thus, more tests are expected when infection is more prevalent. The national database does not contain reliable data on individuals’ symptoms or on their reasons for performing PCR testing. We attempted to control for individuals who tend to have more PCR tests (e.g., healthcare workers or people in nursing homes) by including past PCR tests (0, 1 or 2+) as a covariate in the model. Figure S2 in the Supplementary Appendix compares the number of PCR tests performed during the study period stratified by the number of PCR tests taken in the past. The association between the variables is evident, showing that including the number of past PCR tests can eliminate part of the bias. Except for those fully vaccinated during January, the numbers of PCR tests taken in the different vaccination periods are quite similar within each age group and past PCR testing level. This suggests that detection bias is not the main reason for the observed waning in Figure 3, and it is probably not a dominant factor. Moreover, all hospitalized patients with severe respiratory disease are tested for SARS-CoV-2 by PCR, and detection bias is minimal in this population. Thus, waning immunity against severe disease shown in Figure 4 strongly suggests that detection bias cannot fully explain the exhibited decrease in protection against infection.

This paper does not quantify the contribution to vaccine breakthrough due to the change in dominant variant from Alpha to Delta or the extent of waning in the months immediately following vaccination (when the prevalence was extremely low in Israel). Those two factors could be as substantial as the waning effect but are beyond the scope of this work. Our analysis reveals the clear effect of waning on top of those two factors that should be separately explored in future studies.

Understanding the extent of waning immunity is critical for policy making, especially in regard to vaccination strategies. The results presented in this paper were the basis of the decision by the Israeli Ministry of Health to give a third dose of COVID-19 vaccine to people aged 60 or over who had been vaccinated at least five months previously, starting July 30, 2021. It also suggests the need to closely follow the effects of waning immunity and inform policy makers facing the decision of administering booster vaccinations worldwide. The 3^rd^ dose campaign in Israel will be closely monitored in order to quantify the effectiveness of a booster dose in restoring protection that has waned over time.

## Data Availability

Aggregated data are given in the supplementary information. Personal data cannot be shared due to
privacy.

## Ethics statement

The study was approved by the Institutional Review Board of the Sheba Medical Center. Helsinki approval number: SMC-8228-21.

## Data sharing

Aggregated data are given in the supplementary information. Personal data cannot be shared due to privacy.

## Acknowledgments

The authors would like to thank Boaz Lev, Ami Mizrahi, Ronen Fluss, Sarah Goldberg, Geert Molenberghs, Rami Yaari, and Arnona Ziv, for fruitful discussions.

## Supplementary Appendix

**Table S1:** Number of persons, person-days and events for the different second dose vaccination periods from July 11, 2021 to July 31, 2021.

| Time period | Age category | Person-days | Positive SARS-CoV-2 tests | Severe COVID-19 |
| --- | --- | --- | --- | --- |
| Jan, 16-31 | 16-39 | 125,587 | 445 | 1 |
| Jan, 16-31 | 40-59 | 242,866 | 929 | 7 |
| Jan, 16-31 | 60+ | 705,313 | 2,266 | 207 |
| Feb, 1-15 | 16-39 | 195,580 | 725 | 1 |
| Feb, 1-15 | 40-59 | 417,618 | 1,406 | 5 |
| Feb, 1-15 | 60+ | 358,020 | 942 | 89 |
| Feb, 16-28 | 16-39 | 352,247 | 1,021 | 0 |
| Feb, 16-28 | 40-59 | 327,766 | 1,016 | 3 |
| Feb, 16-28 | 60+ | 66,931 | 145 | 9 |
| Mar, 1-15 | 16-39 | 549,089 | 1,426 | 0 |
| Mar, 1-15 | 40-59 | 208,072 | 535 | 3 |
| Mar, 1-15 | 60+ | 61,814 | 104 | 13 |
| Mar, 16-31 | 16-39 | 496,455 | 937 | 0 |
| Mar, 16-31 | 40-59 | 190,200 | 344 | 0 |
| Mar, 16-31 | 60+ | 62,277 | 102 | 5 |
| Apr | 16-39 | 217,622 | 294 | 0 |
| Apr | 40-59 | 78,249 | 112 | 1 |
| Apr | 60+ | 29,125 | 37 | 4 |
| May | 16-39 | 67,281 | 96 | 0 |
| May | 40-59 | 22,240 | 30 | 0 |
| May | 60+ | 10,893 | 15 | 0 |

**Figure S1:**
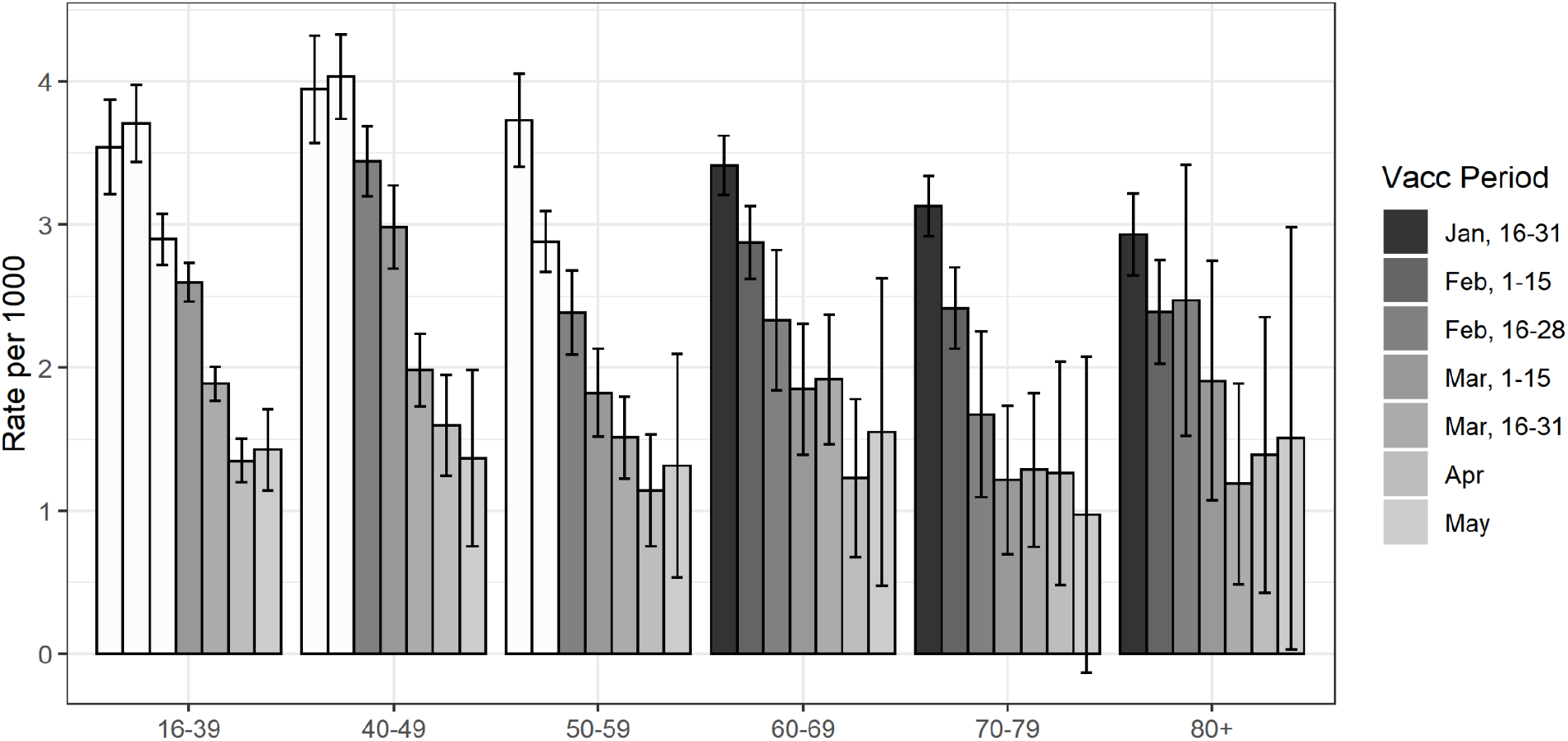
Rate of documented SARS-CoV-2 infection (per 1,000 persons) from July 11, 2021 to July 31, 2021, stratified by period of second dose of COVID-19 vaccine and a finer age grouping. White bars represent periods at which only persons at higher risk were allowed to receive vaccination.

**Table S2:**
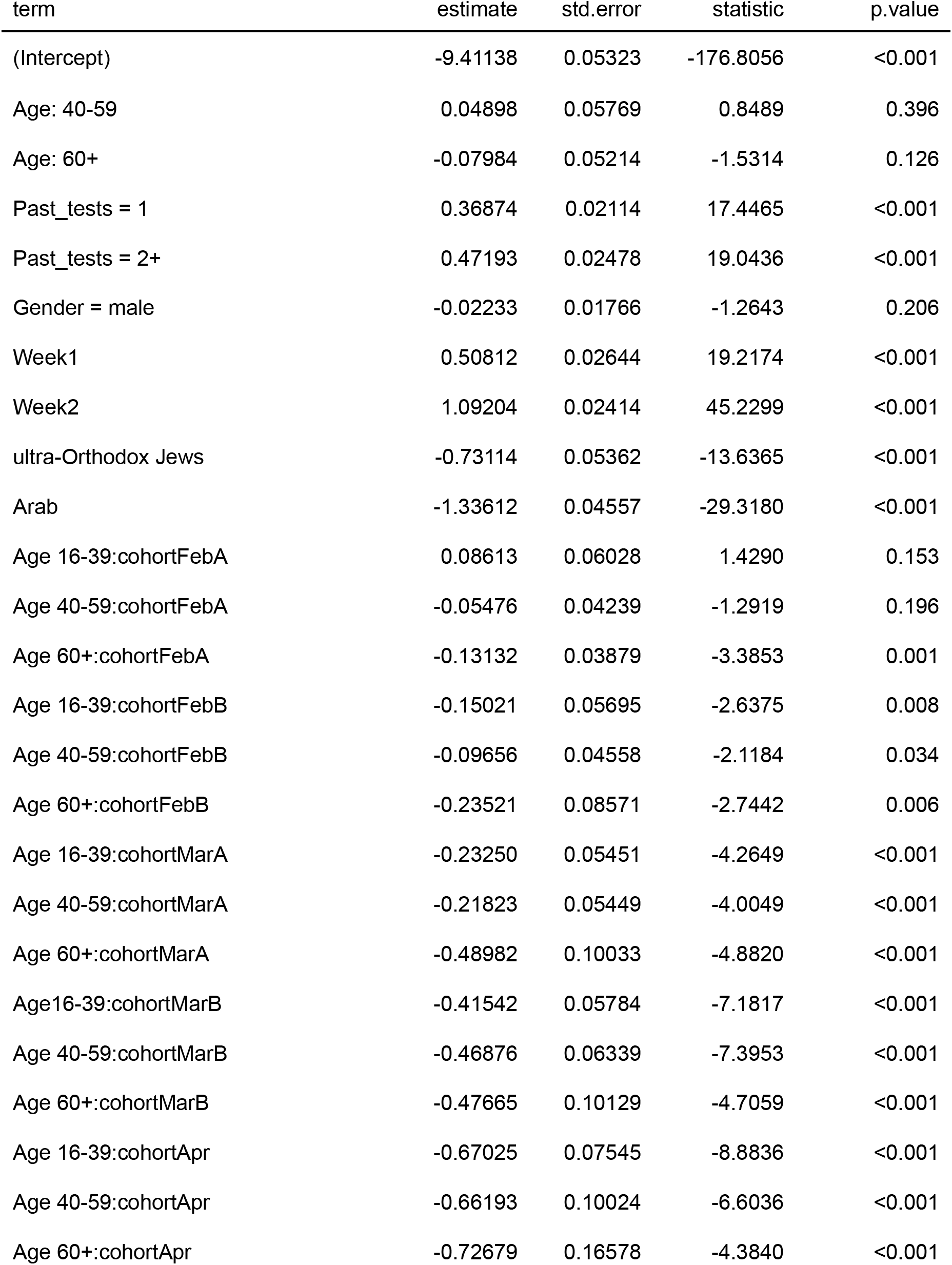
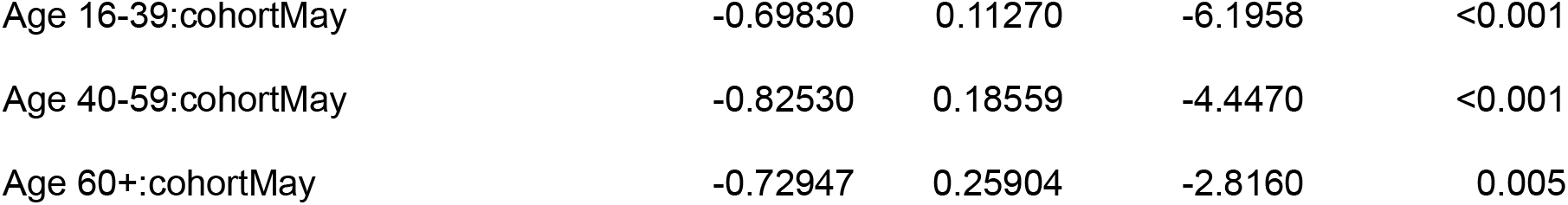
Poisson regression results for documented SARS-CoV-2 infection by PCR. A and B indicate the first and second halves of the corresponding months.

| term | estimate | std.error | statistic | p.value |
| --- | --- | --- | --- | --- |
| (Intercept) | -9.41138 | 0.05323 | -176.8056 | <0.001 |
| Age: 40-59 | 0.04898 | 0.05769 | 0.8489 | 0.396 |
| Age: 60+ | -0.07984 | 0.05214 | -1.5314 | 0.126 |
| Past_tests = 1 | 0.36874 | 0.02114 | 17.4465 | <0.001 |
| Past_tests = 2+ | 0.47193 | 0.02478 | 19.0436 | <0.001 |
| Gender = male | -0.02233 | 0.01766 | -1.2643 | 0.206 |
| Week1 | 0.50812 | 0.02644 | 19.2174 | <0.001 |
| Week2 | 1.09204 | 0.02414 | 45.2299 | <0.001 |
| ultra-Orthodox Jews | -0.73114 | 0.05362 | -13.6365 | <0.001 |
| Arab | -1.33612 | 0.04557 | -29.3180 | <0.001 |
| Age 16-39:cohortFebA | 0.08613 | 0.06028 | 1.4290 | 0.153 |
| Age 40-59:cohortFebA | -0.05476 | 0.04239 | -1.2919 | 0.196 |
| Age 60+:cohortFebA | -0.13132 | 0.03879 | -3.3853 | 0.001 |
| Age 16-39:cohortFebB | -0.15021 | 0.05695 | -2.6375 | 0.008 |
| Age 40-59:cohortFebB | -0.09656 | 0.04558 | -2.1184 | 0.034 |
| Age 60+:cohortFebB | -0.23521 | 0.08571 | -2.7442 | 0.006 |
| Age 16-39:cohortMarA | -0.23250 | 0.05451 | -4.2649 | <0.001 |
| Age 40-59:cohortMarA | -0.21823 | 0.05449 | -4.0049 | <0.001 |
| Age 60+:cohortMarA | -0.48982 | 0.10033 | -4.8820 | <0.001 |
| Age16-39:cohortMarB | -0.41542 | 0.05784 | -7.1817 | <0.001 |
| Age 40-59:cohortMarB | -0.46876 | 0.06339 | -7.3953 | <0.001 |
| Age 60+:cohortMarB | -0.47665 | 0.10129 | -4.7059 | <0.001 |
| Age 16-39:cohortApr | -0.67025 | 0.07545 | -8.8836 | <0.001 |
| Age 40-59:cohortApr | -0.66193 | 0.10024 | -6.6036 | <0.001 |
| Age 60+:cohortApr | -0.72679 | 0.16578 | -4.3840 | <0.001 |
| Age 16-39:cohortMay | -0.69830 | 0.11270 | -6.1958 | <0.001 |
| Age 40-59:cohortMay | -0.82530 | 0.18559 | -4.4470 | <0.001 |
| Age 60+:cohortMay | -0.72947 | 0.25904 | -2.8160 | 0.005 |

**Table S3:** Poisson regression results for severe COVID-19. A and B indicate the first and second halves of the corresponding months.

| term | estimate | std.error | statistic | p.value |
| --- | --- | --- | --- | --- |
| (Intercept) | -15.02352 | 0.4106 | -36.58897 | <0.001 |
| Age: 60+ | 2.49480 | 0.3851 | 6.47818 | <0.001 |
| Past_tests = 1 | 0.26677 | 0.1522 | 1.75284 | 0.08 |
| Past_tests = 2+ | 1.26406 | 0.1268 | 9.97142 | <0.001 |
| Gender = male | 0.63268 | 0.1113 | 5.68351 | <0.001 |
| Week1 | 0.53596 | 0.1698 | 3.15708 | 0.002 |
| Week2 | 1.25016 | 0.1529 | 8.17429 | <0.001 |
| Orthodox Jews | -0.01861 | 0.2734 | -0.06807 | 0.946 |
| Israeli Arab | -0.40339 | 0.2453 | -1.64424 | 0.1 |
| Age 40-59:cohortFeb | -0.79237 | 0.5182 | -1.52921 | 0.126 |
| Age 60+:cohortFeb | -0.14240 | 0.1236 | -1.15253 | 0.249 |
| Age 40-59:cohortMar | -1.03054 | 0.6914 | -1.49052 | 0.136 |
| Age 60+:cohortMar | -0.50925 | 0.2479 | -2.05458 | 0.04 |

### Supplementary Analysis 1 - Comparison to the unvaccinated group

We studied the protection of fully vaccinated individuals compared to that of unvaccinated individuals without evidence of prior infection with SARS-CoV-2 by PCR at the start of the study. As unvaccinated uninfected individuals were not part of the database (unless they performed a SARS-CoV-2 PCR test in the past), data on all residents of Israel stratified by age, gender, and demographic group were obtained from the Israel Central Bureau of Statistics, and were merged with the Ministry of Health database into a unified dataset of all residents of Israel (n=9,395,923): unvaccinated, vaccinated, and previously infected individuals.

The rates of documented SARS-CoV-2 infection and severe COVID-19 were converted into estimates of protection by comparing them to the rates of the unvaccinated group. Specifically, we repeated the analysis described in the Methods section on the unified data, including the unvaccinated group as an additional “vaccination period” in the model (reference group). We then compared the protection of vaccinated individuals in different periods adjusting for age, week, past PCR tests, demographic group, and gender. As in previous analyses, we included an interaction term between age and period of vaccination in the model in order to calculate age-specific effectiveness.

**Table S4.** Vaccine effectiveness against documented SARS-CoV-2 infection and severe COVID-19 [95% CI] compared to the unvaccinated cohort for the different age groups, adjusted for week of infection, past PCR testing, demographic group, and gender. For severe COVID-19, estimates are not provided for the lowest age group and for the last vaccination periods due to very low case numbers. A and B indicate the first and second halves of the corresponding months.

| OUTCOME = Positive SARS-CoV-2 PCR test |  |  |  |  |  |  |  |
| --- | --- | --- | --- | --- | --- | --- | --- |
| Age | JanB | FebA | FebB | MarA | MarB | Apr | May |
| 16-39 | 50% [45, 55] | 47% [42, 52] | 58% [55, 62] | 62% [59, 64] | 68% [65, 70] | 74% [71, 77] | 73% [67, 78] |
| 40-59 | 58% [54, 62] | 61% [58, 65] | 63% [59, 66] | 67% [63, 70] | 74% [70, 77] | 78% [73, 82] | 80% [71, 86] |
| 60+ | 57% [52, 62] | 63% [57, 67] | 65% [57, 71] | 73% [66, 78] | 72% [64, 77] | 73% [63, 81] | 75% [58, 85] |
| OUTCOME = Severe COVID-19 |  |  |  |  |  |  |  |
| Age | Jan | Feb | Mar |  |  |  |  |
| 40-59 | 94% [87, 97] | 98% [95, 99] | 98% [94, 99] |  |  |  |  |
| 60+ | 86% [82, 90] | 88% [84, 91] | 91% [85, 95] |  |  |  |  |

**Figure S2.**
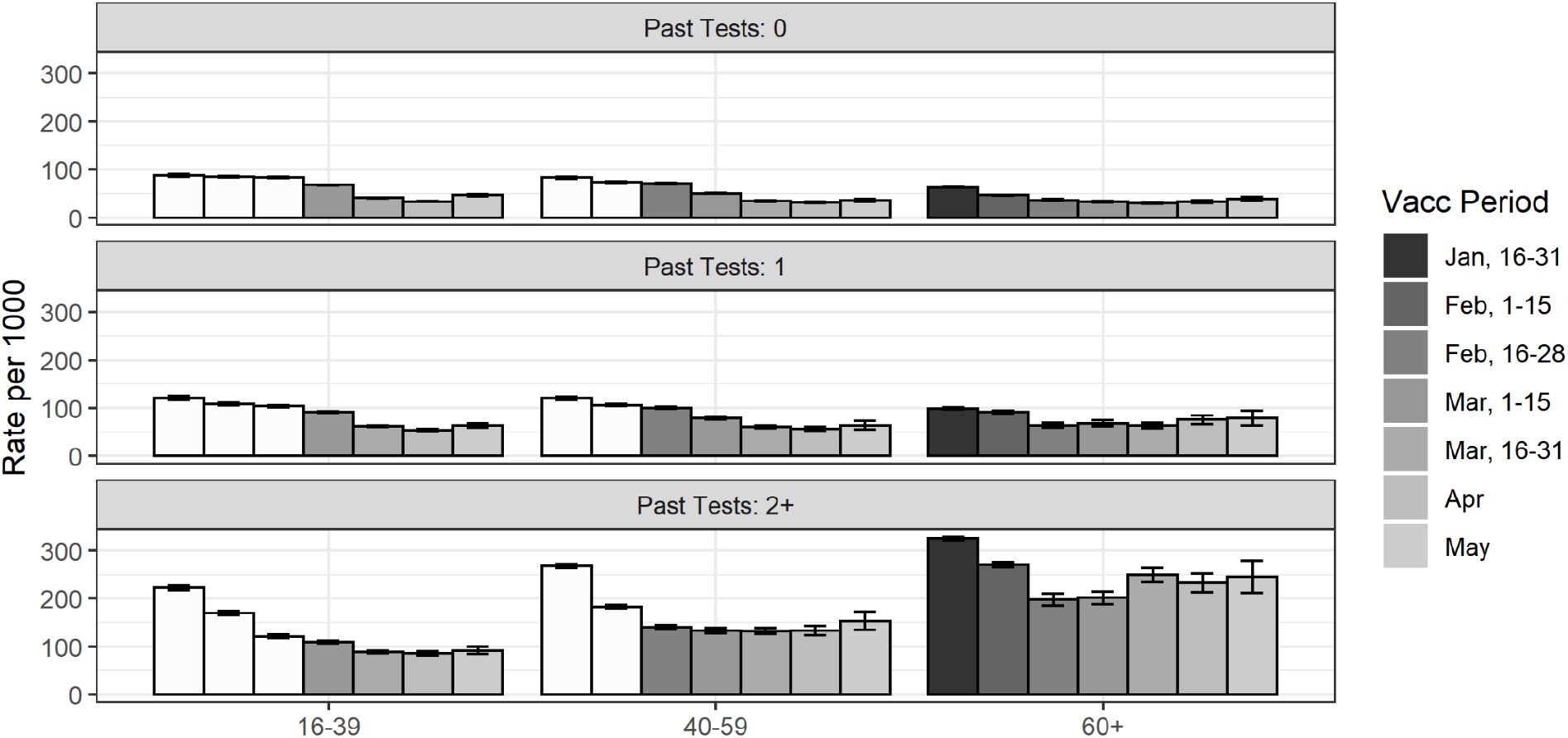
Number of SARS-CoV_2 PCR tests performed from July 11, 2021 to July 31,2021, grouped by vaccination period and age group, and stratified by past PCR tests. White bars represent periods during which only persons at higher risk were allowed to receive the vaccine.

